# Open-source 3D Printable Forearm Crutch

**DOI:** 10.1101/2024.11.12.24317179

**Authors:** Maryam Mottaghi, Morgan Woods, Laura Danier, Anita So, Jacob M. Reeves, Joshua M. Pearce

## Abstract

Although there has been considerable progress in distributed manufacturing of open-source designs for mobility aids, there is a notable lack of affordable, open-source crutches. Crutches are a vital tool for many individuals with mobility impairments, yet the high costs limit accessibility. Even more, they are in short supply in regions undergoing conflict. The goal of this study is to address this need by leveraging the principles of free and open-source hardware and the capabilities of digital distributed manufacturing to create a low-cost, functional crutch that can be easily produced and customized locally using inexpensive desktop 3D printers. All the design files are open-source, and the design process incorporated load-bearing tests using a hydraulic actuator under static loading conditions to meet the ISO 11334-1:2007 standard for walking aids. The open-source forearm crutch developed in this study not only surpasses the requirements of the ISO method for load capacity (1,516.3 ± 169.9 N, which is 51.6% percent above needs), weighs a fraction of comparable commercial systems (0.612 kg or 27%% of proprietary devices), and is customizable, but also offers a highly cost-effective solution; costing CAD $36 in material, which is less than all equivalent crutches on the open market. If recycled plastic is used, the material cost of the crutch could be further reduced to under CAD $13, making it much more accessible.

## 1. Introduction

Mobility-related disabilities currently impact over 10% of the adult population globally [1,2], with projections only expected to rise dramatically as the population ages [3]. Conditions such as arthritis, chronic back pain, and injuries from accidents contribute to the increased need for mobility aids, particularly among older adults [3]. By 2060, nearly a quarter of the U.S. population will be 65 years or older [4]. Addressing the related challenges expected to be faced by those with mobility impairments requires attention. These challenges are not only physical but also economic, as many individuals, especially in less developed regions or those living in poverty, struggle to afford the adaptive mobility aids they require for daily living [5].

Commercial mobility aids, such as crutches, canes, walkers, and wheelchairs, are available on the market, but acquiring them can be challenging for many individuals due to their high costs [6]. For instance, proprietary forearm crutches can range in price from CAD$36.66 to CAD$315.00 making them unaffordable for individuals on restricted incomes or without extended health coverage (Table 1). Individuals with disabilities face a higher risk of unemployment, which in turn increases their likelihood of living in poverty [7]. This economic vulnerability makes it even more difficult for them to afford essential mobility aids [7].

**Table 1.** List of price and source of commercial crutches.

| **Name of the Product** | **Price (CAD\$)** | **Source (Date accessed October 16, 2024)** |
| --- | --- | --- |
| Folding Crutch | \$36.66 | https://www.amazon.ca/Disabled-Portable-Adjustable-Underarm-Telescopic/dp/B08J2JCDGB/ |
| Forearm Crutches | \$47.70 | https://www.walmart.com/ip/Medline-Walking-Forearm-Crutches-Lightweight-Aluminum-250lb-Weight-Capacity-1-Pair/22790105 |
| PEPE - Crutches | \$59.99 | https://www.amazon.ca/PEPE-Crutches-Forearm-Aluminum-Adjustable/dp/B07DPQ2S2F/ |
| Forearm Crutches | \$99.99 | https://www.amazon.ca/HEALTHBAZAAR-Crutches-Fashionable-Ergonomic-Handgrips/dp/B0CPHSWZB8/ |
| ORTONYX Forearm Crutches with Adjustable Support | \$161.76 | https://www.amazon.ca/ORTONYX-Adjustable-Ergonomic-Comfortable-Lightweight/dp/B07C1QXN12/ |
| Ergonomic Forearm Crutches | \$315.00 | https://www.amazon.ca/Ergobaum®-Ergoactives-Ergonomic-Forearm-Crutches/dp/B00LGZ43E2/ |

Additionally, this lack of access to essential mobility aids not only makes individuals with disabilities dependent on others but also increases social inequalities, as those without the financial means are further impacted.

Digital distributed manufacturing offers a promising method for addressing these challenges [8–10]. Additive manufacturing, which is facilitated by Computer Numerical Control (CNC) tools like 3D printers, has the potential to drastically reduce the cost of consumer goods, including adaptive mobility aids [11]. The transformation of 3D printing technology from a tool for creating prototypes to a widely accessible method for producing usable goods has opened up new possibilities for manufacturing affordable products locally [12,13]. This technology has been adopted by local businesses [11][14,15], chain stores [16], makerspaces [17–19], fablabs [20], and even public libraries[21–23], which makes production more accessible and allows for the customization of products to fit individual needs [24].

The proliferation of free and open-source hardware (FOSH) has further expedited this revolution [25]. The open-source movement, which began in software development, has evolved into a powerful framework for hardware innovation [26]. The release of self-replicating rapid prototyper (RepRap) designs allowed 3D printers to produce many of their own components, which significantly reduced their costs [27–29]. Today, there are millions of 3D printable FOSH designs available [30], which provide consumers with the opportunity to create customized products at a fraction of the cost of commercial products [31].

Adaptive aids are particularly well-suited to benefit from the open-source model, because of the advantages of customization and relatively high-markup on such products [32]. For example, a low-cost open-source walker has already been developed [11]. This open-source walker is constructed from readily available materials and 3D printed joints which together meet the weight requirements for most users up to 187.1 ± 29.3 kg while reducing walker costs and mass compared to commercial alternatives [11].

Despite the advances in open-source mobility aids, there remains a critical gap in the availability of affordable, open-source forearm crutches. Forearm crutches are a vital tool for many individuals with mobility impairments, yet the high cost and lack of customization options in commercial products can be challenging. The development of an open-source crutch that is both affordable and customizable could transform the lives of countless individuals and offer them greater independence in mobility. This paper aims to address this need by leveraging the principles of FOSH and the capabilities of digital distributed manufacturing to create a low-cost, forearm crutch that can be easily produced and customized locally. Specifically, low-cost open-source desktop 3D printing is used to manufacture bespoke components. The design process incorporated repetitive testing to ensure that the crutch could meet standardized load-bearing requirements. For load-bearing tests, mechanical tests were performed using an MTS hydraulic actuator with a capacity of 250 kN under static loading conditions. The crutch was subjected to the static load capacity requirements outlined in the ISO 11334-1:2007 standard for walking aids. This standard specifies that crutches must support a load of 1000 N ± 2% which simulates a user mass of 100 kg, without breaking or deforming for a specified duration of 10 seconds.

## 2. Materials and Methods

### 2.1. Crutch Design

Following the evaluation of various commercially available crutches, an initial design concept for an open-source forearm crutch was designed in Onshape CAD software (Onshape 1.157, Cambridge, MA, USA) [33] using 3D printed joints in combination with commercially available hardwood dowels. Solid, cylindrical hardwood dowels were selected as the primary structural components due to their sustainability, ease of availability in standard sizes across hardware stores in North America, and compatibility with the 3D printed joints. Hardwood was chosen not only for its renewable and biodegradable properties but also for its potential to be recycled and composted. Furthermore, a study comparing the carbon footprint of wood and aluminum—a common structural material for mobility aids—revealed that wood generates approximately one-fourth of the carbon emissions compared with aluminum in the context of window frames [34]. It is important to note that the strength of the crutch will vary depending on the specific type of wood used, as hardwood can encompass a variety of types such as basswood, beech, maple, or oak, and softwoods include types such as cedar, pine, or spruce [35]. For the present investigation hardwood dowel was chosen, though the specific source is commercially unspecified.

This open-source forearm crutch design incorporates several features inspired by existing commercially available devices. Notably, the handle and cuff are integrated into a single 3D printed part (Figure 1). In accordance with the ISO 11334-1:2007 standard, the handle is ergonomically shaped to align with the handgrip support line to ensure that the user’s hand and wrist is in a comfortable position during use. The arm section length, located above the rear handgrip reference point, features a non-horizontal forearm support (cuff) that secures the forearm in position and prevents lateral movement. The arm section length (a), cuff internal depth (x), width (y) (Figure 2a), and height (z) (Figure 2b) can be tailored to provide comfortable support for the forearm (Figure 2c). The diameter of the cuff is configured as a variable in the CAD file to allow for easy customization. Since wood is a renewable material that can be easily recycled and composted, a standardized wooden dowel with a solid, circular cross-section and diameter of 22.3 mm (7/8” standard hardwood dowel) was selected for the leg section [36]. The dowels are available in standard sizes in North America and could be slotted directly into the solid 3D printed forearm handle and cuff. To better facilitate the connection between the leg section and the 3D printed handle, a TPU washer (Figure 3a) was placed between the wooden dowel and 3D printed components. This washer offered both shock absorption and tolerance between the potentially unparallel surfaces of the cut dowel surface and 3D print to ensure the load was more evenly transferred through the dowel to the 3D printed handle. Similarly, a TPU handle grip (Figure 3b) was modeled and 3D printed to slide onto the handle to further promote user comfort.

**Figure 1.**
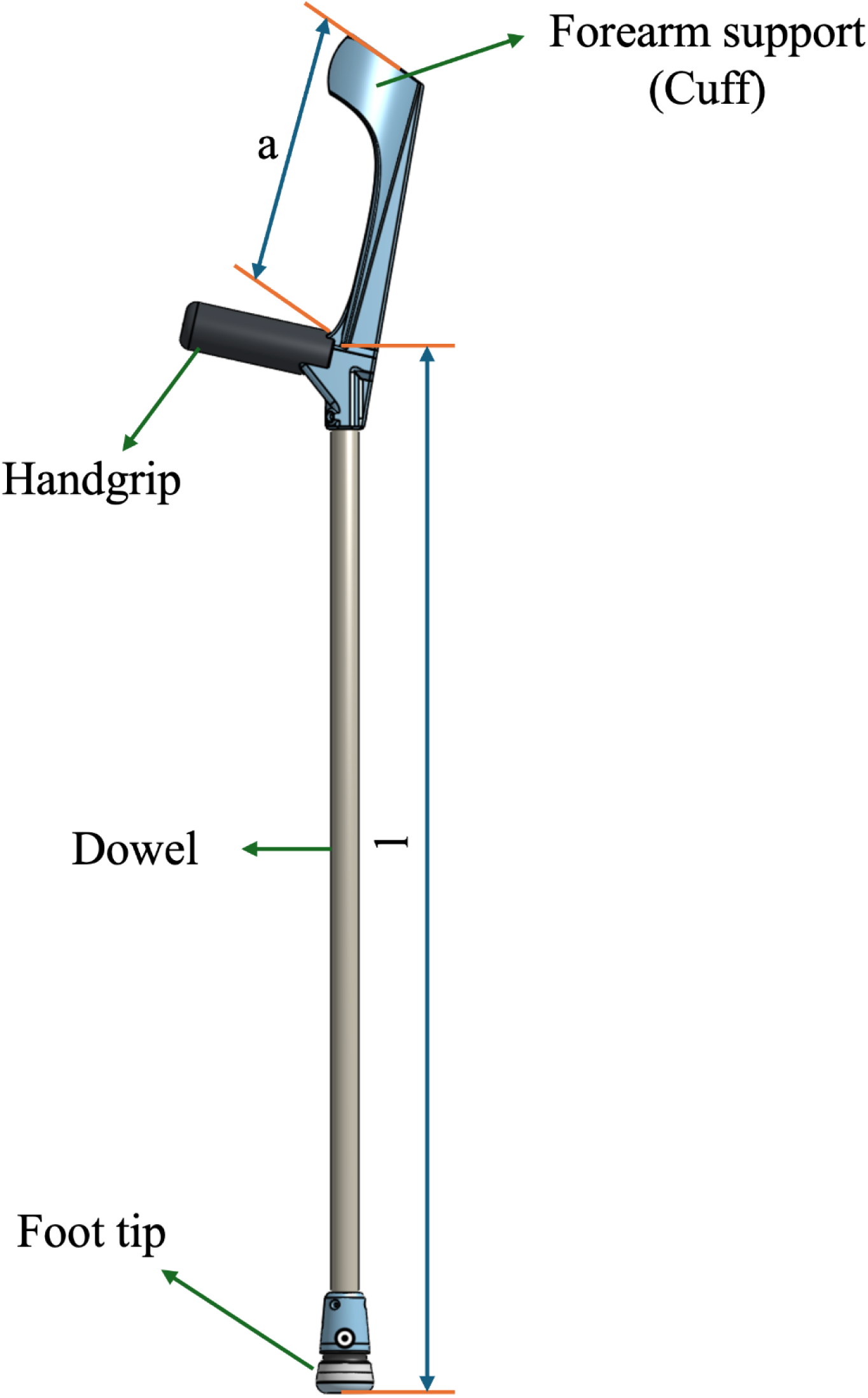
Dimensional measurements of assembled crutch. a) length of forearm 227 mm, l) length of leg section 794 mm.

**Figure 2.**
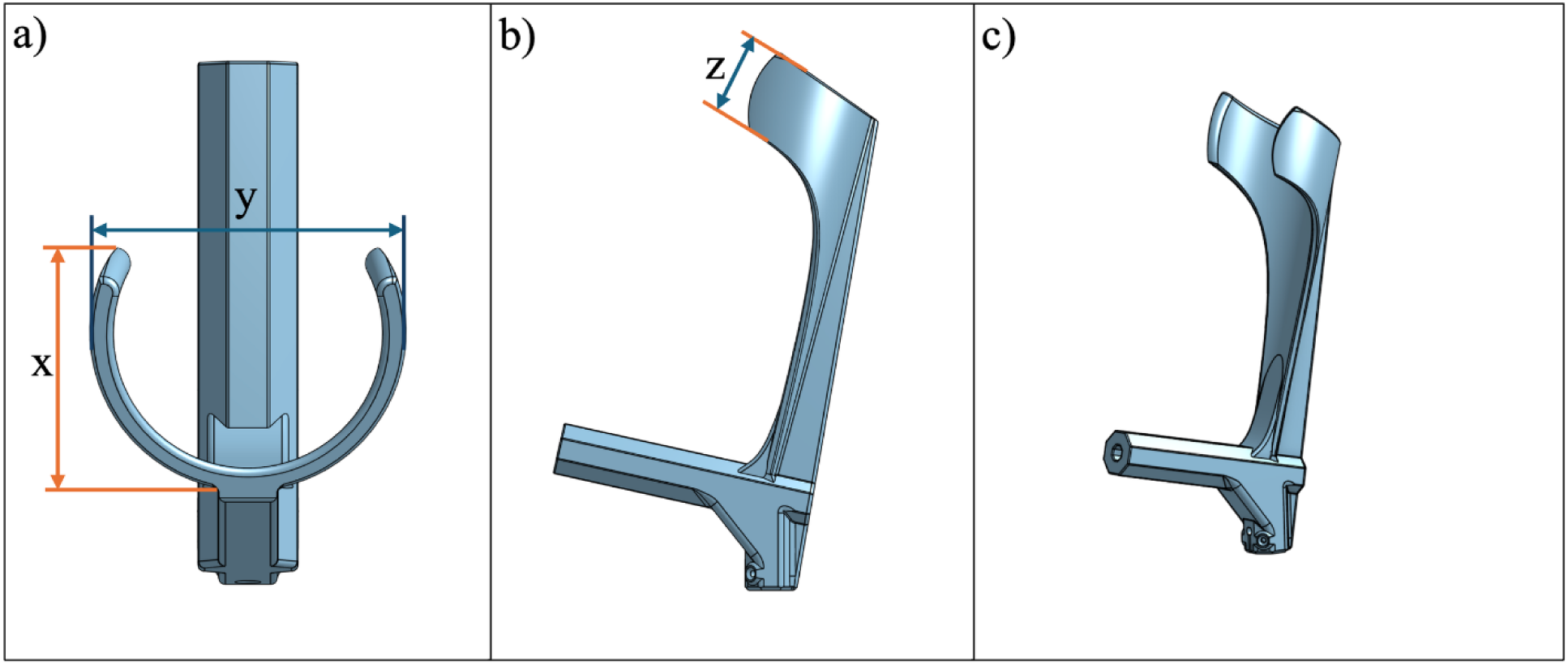
Forearm design a) top view, featuring (x) cuff depth and (y) cuff width; b) right view; c) isometric view.

**Figure 3.**
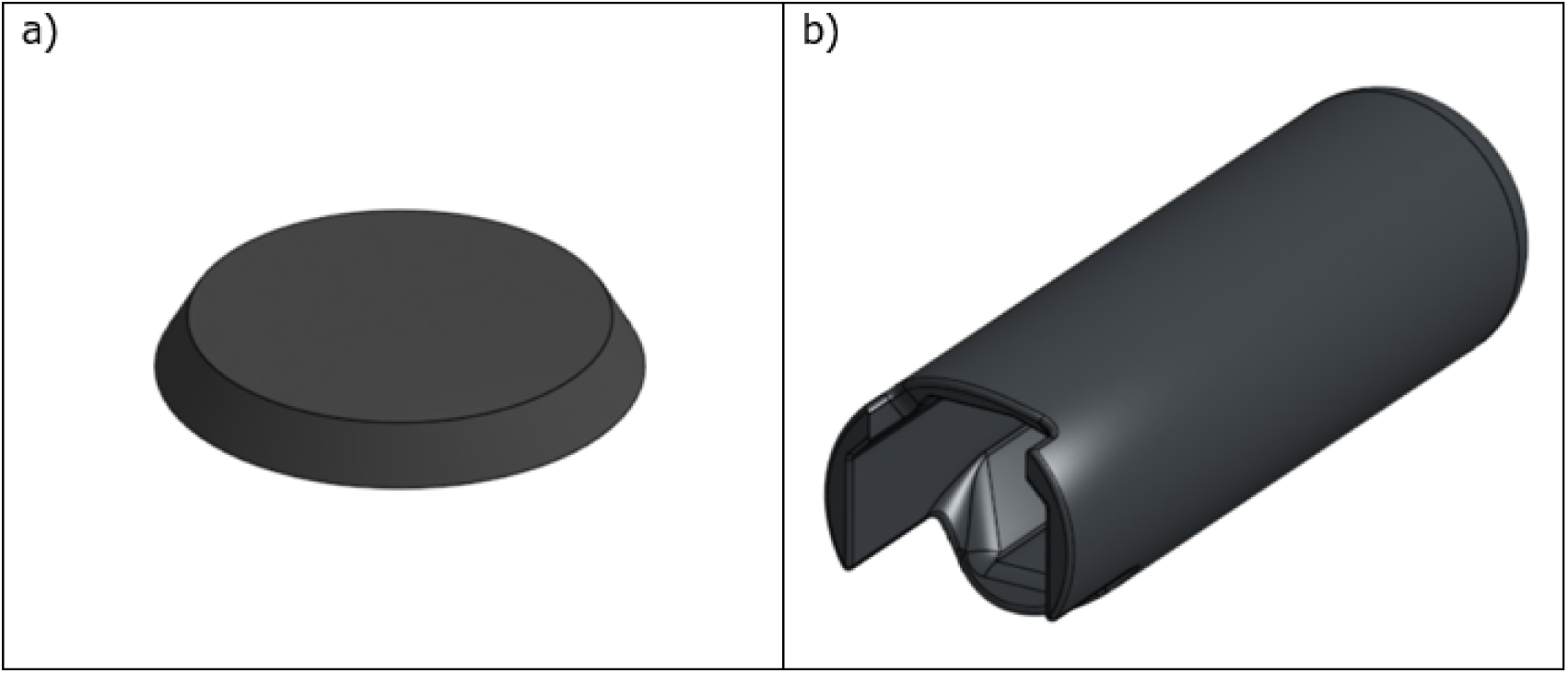
Design of a) washer (TPU-85A), b) handle grip (TPU-85A).

The leg section, located below the rear handgrip reference point, ends with a tip designed to ensure stable contact with the ground. The leg section length (l), as was the arm section length (a), are adjustable and can be customized to accommodate users of various heights (Figure 1).The foot tip (Figure 4) is constructed from the assembly of a foot cushion (Figure 4a), foot base (Figure 4b), foot living joint (Figure 4c), and ankle body (Figure 4d). The assembly of the foot tip is shown in Figure 4e. All 3D printed connections are secured to the wooden dowel using #6 x 5/8” flat head wood screws to ensure stability.

**Figure 4.**
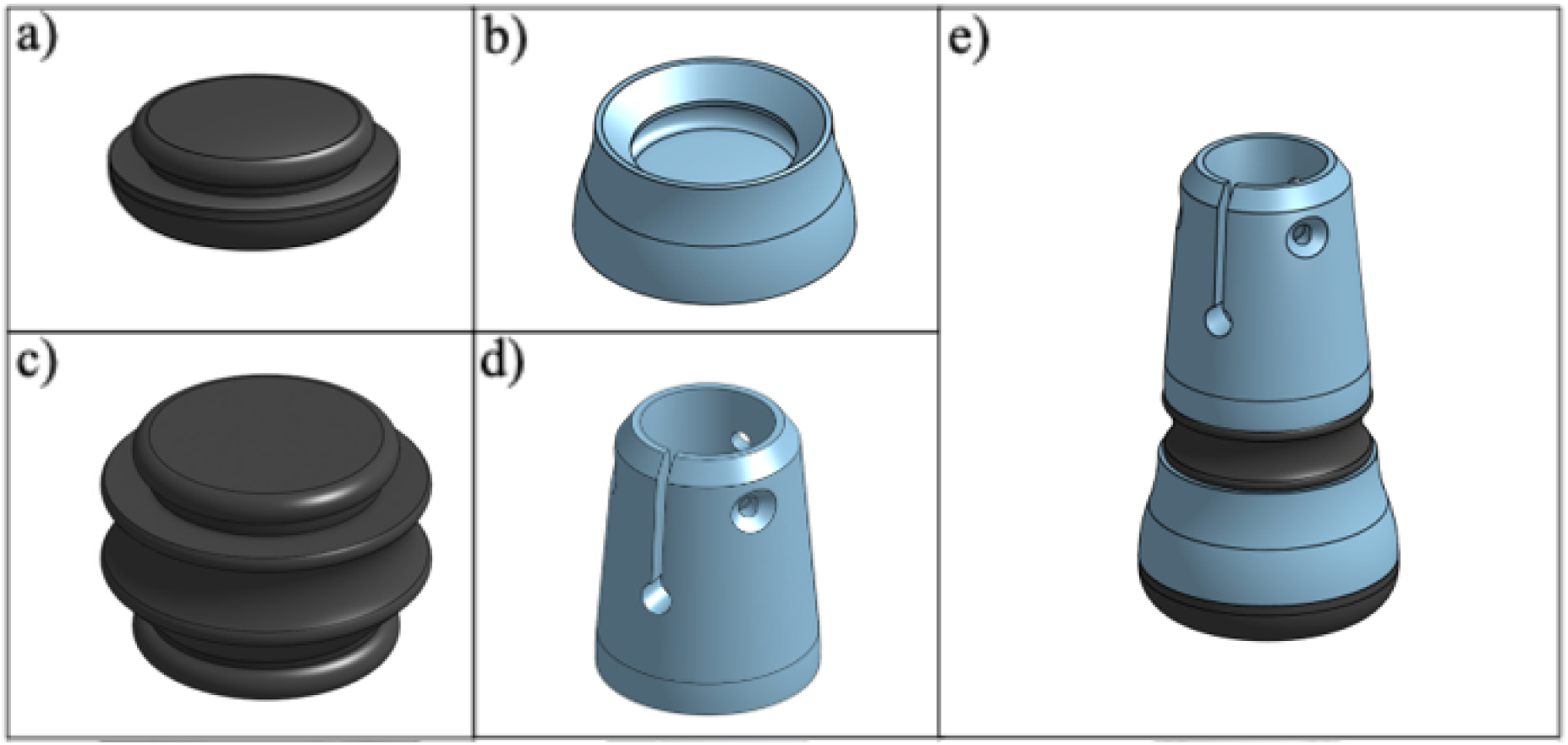
Design of a) foot cushion (TPU-85A), b) foot base (PETG), c) foot living joint (TPU-85A), d) ankle body (PETG), and e) assembled foot tip.

### 2.2. Overview of 3D Printing and Wood Dowel Part Sizing

All 3D printed components were produced using Polymaker PETG filament [37], or Ninja Flex TPU 85A [38], with printing performed on open-source RepRap-class 3D printers, either the LulzBot TAZ Workhorse [39], or Prusa i3 Mk3S (Prusa, Prague, Czech Republic) [40]. Each of which was equipped with a 0.6 mm nozzle. An index of the 3D printed parts, including their names and quantities, are detailed in Table 2. The slicing parameters used for both PETG and TPU are outlined in Table 3.

**Table 2.** Bill of materials for 3D printed components.

| Name | Number of Parts |
| --- | --- |
| One-piece forearm | 1 |
| Handle Grip | 1 |
| Washer | 2 |
| Foot cushion | 1 |
| Foot | 1 |
| Foot living joint | 1 |
| Ankle body | 1 |

**Table 3.** Slicing parameters for PETG and TPU 85A filaments.

| Slicing Parameter | PETG Value | TPU 85A Value |
| --- | --- | --- |
| Layer Height | 0.6 mm | 0.15 |
| Wall Count | 4 | 2 |
| Infill Density | 99% | 30% |
| Infill Pattern | Gyroid | Gyroid |
| Printing Temperature | 230 °C | 238 °C |
| Bed Temperature | 85 °C | 50 °C |

To address the complex geometries of the parts, each component was strategically oriented on the print bed to reduce alignment with potential fracture planes and to decrease filament usage by reducing the need for support structures. Support structures are necessary for some overhangs, as illustrated in Figure 5.

**Figure 5.**
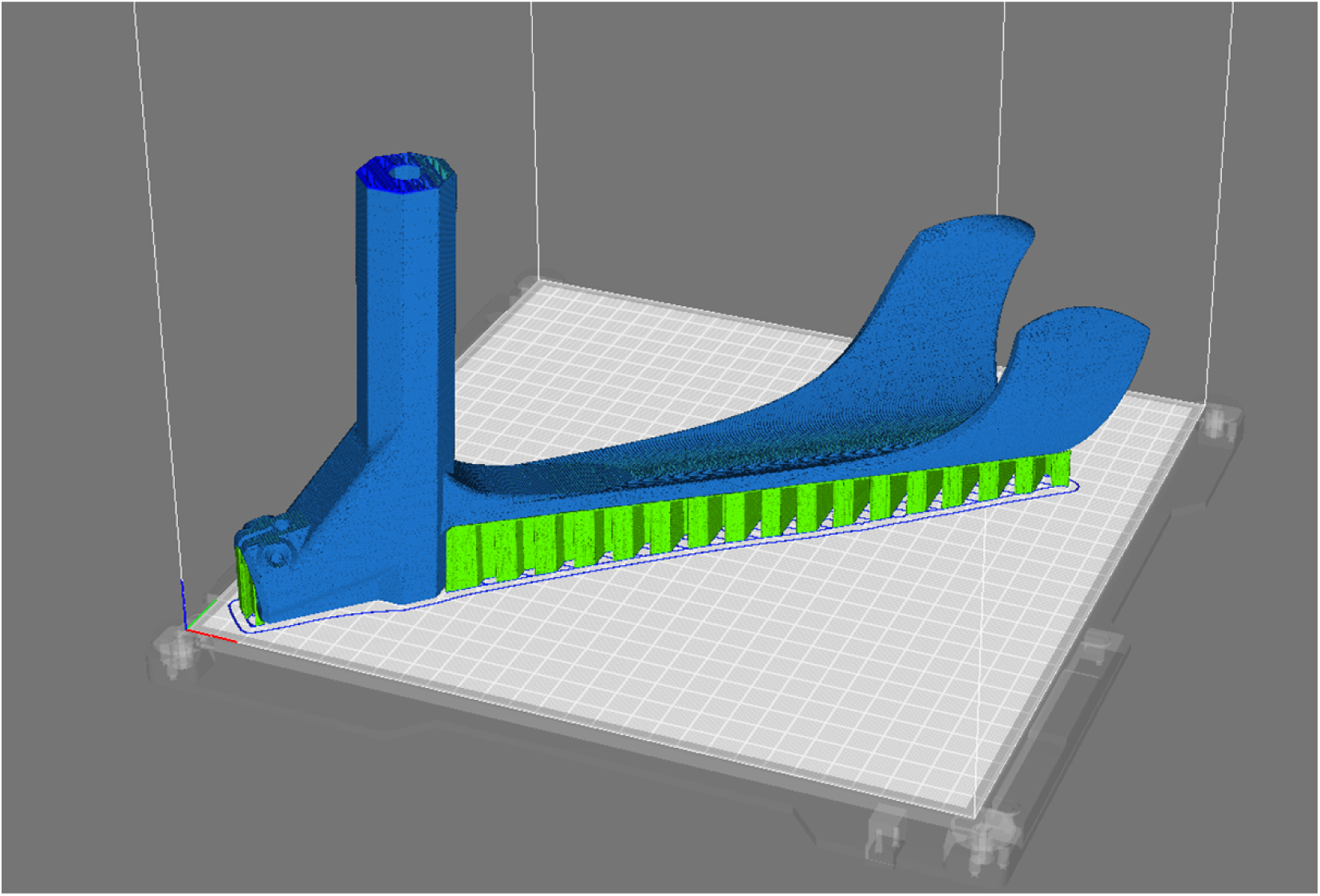
Orientation of the 3D printed one-piece forearm (blue) on the print bed with support locations (green)

The height of the forearm crutch is indicated from the handgrip to the floor, with an adjustable range of 740 to 890 mm [41]. To ensure the crutch design targeted the worst-case loading scenario with a higher chance of buckling and more severe failure, the wooden dowel was cut to 794 mm, which resulted in a distance from the handgrip to the floor of 890 mm. This ensured the wrist location matches the tallest crutch specifications. Once the tallest case was validated, the height could be further customized based on the length between the user’s hand and the ground with certainty of loading capability.

### 2.3. Mechanical Testing

To ensure the crutch can safely withstand regular use, compressive mechanical testing was conducted to determine the load required to cause failure. The methods used were adapted from the “Static loading” criteria outlined in Section 5.6 of ISO 11334-1:2007. According to this standard, crutches must support a vertical load of 1000 N ± 2% without breaking or deforming under a user mass of 100 kg for a specified duration of 10 seconds. To provide additional insights into the failure mode of the design and observe any potential weak points, each crutch was subsequently loaded to its breaking point after the 1000 N static loading criteria was achieved.

To simulate the real-world usage of the crutch under static loading conditions, an open-source test jig was developed based on the Section 5.6 of ISO 11334-1:2007 as well as previous investigations [42,43]. The testing jig included a simulated forearm, 3D printed simulated elbow, the testing jig body which interfaced with the hydraulic press, and a 3D printed simulated hand as shown in Figure 6. These components were fabricated and assembled to mimic anatomical structure and movement, and to adhere to the appropriate measurement relationships outlined in ISO 11334-1:2007. The loading force was applied through the testing jig body where it was transferred through the back of the forearm cuff and into the handle through the elbow and wrist joints respectively. These swivel joints and contact points aimed to replicate the user’s natural posture. The swiveling joint that transferred the load to the simulated elbow was positioned just below the cuff, while the simulated forearm rested along the cuff support line and was hinged to the simulated hand. The hand was clamped to the handgrip only at the far end of the handle grip to ensure no restraint or reinforcement could affect the test. The crutch remained free to flex, rotate, and pivot in all directions to simulate realistic motion. The swiveling joint allowed a minimum of 15° of pivot in all directions, while the forearm-hand hinge provided free forward and backward movement, as well as a minimum of 4° of lateral motion.

**Figure 6:**
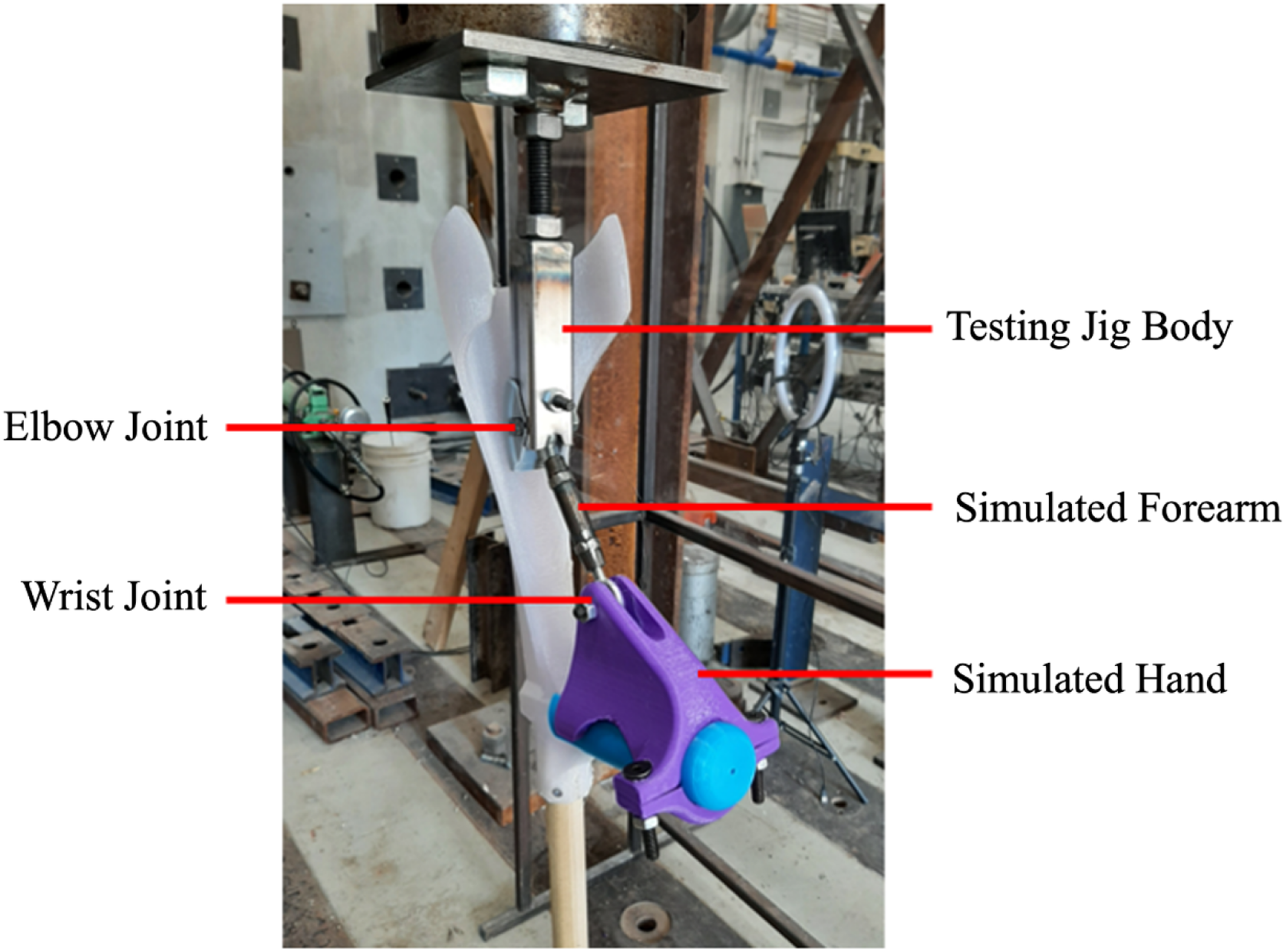
Mechanical testing jig and forearm crutch mounted in the hydraulic press.

Five prototype crutches were tested, and the results were recorded using an MTS hydraulic actuator with a capacity of 250 kN and a stroke length of 150 mm which was capable of operating in both force-controlled and displacement-controlled modes. Force-displacement curves were generated and analyzed to determine the maximum load the crutch could endure before failure. Failure was defined as the point at which the force began to decrease with continued displacement, which indicates the fracturing of either the wooden dowel or the 3D printed components. The stiffness of each crutch was also calculated in Newtons per millimeter (N/mm) by fitting a trendline to the linear region of the force-displacement curves.

The crutch was secured in the universal testing machine, and a vertical force was applied at a controlled rate of 50 mm/min. This rate was selected to ensure the 1000N threshold was achieved in approximately 30 seconds as the standard called for a rate that facilitated 1000N after a minimum of 2 seconds. Throughout testing, no slipping occurred between the foot tip and the ground or between the simulated elbow and testing jig body. The load point was positioned to mimic the force exerted by a user pressing down on the crutch with their hand, in accordance with ISO 11334-1:2007, which specified the requirements and testing methods for walking aids used by individuals with mobility impairments. To improve accuracy, the testing machine was customized with a 3D printed elbow adapter, designed to simulate the user’s elbow. The jig was used to adjust the height of the tester and ensure precise alignment between the machine head and the elbow adapter during testing. The load was gradually increased at 50 mm/min until reaching 1000 N, at which point it was held constant for 10 seconds, before it proceeded at the same rate to failure. This process was repeated across all five of the crutch samples that were fabricated.

## 3. Results and Discussion

All components of the crutch were successfully 3D printed, and support materials were removed using needle nose pliers where applicable. The single piece 3D printed forearm crutch is shown in Figure 7 and the 3D printed foot components are shown in Figure 8 with the supports removed. The fully assembled crutch is depicted in Figure 9a and the crutch is shown in use in Figure 9b and Figure 9c.

**Figure 7.**
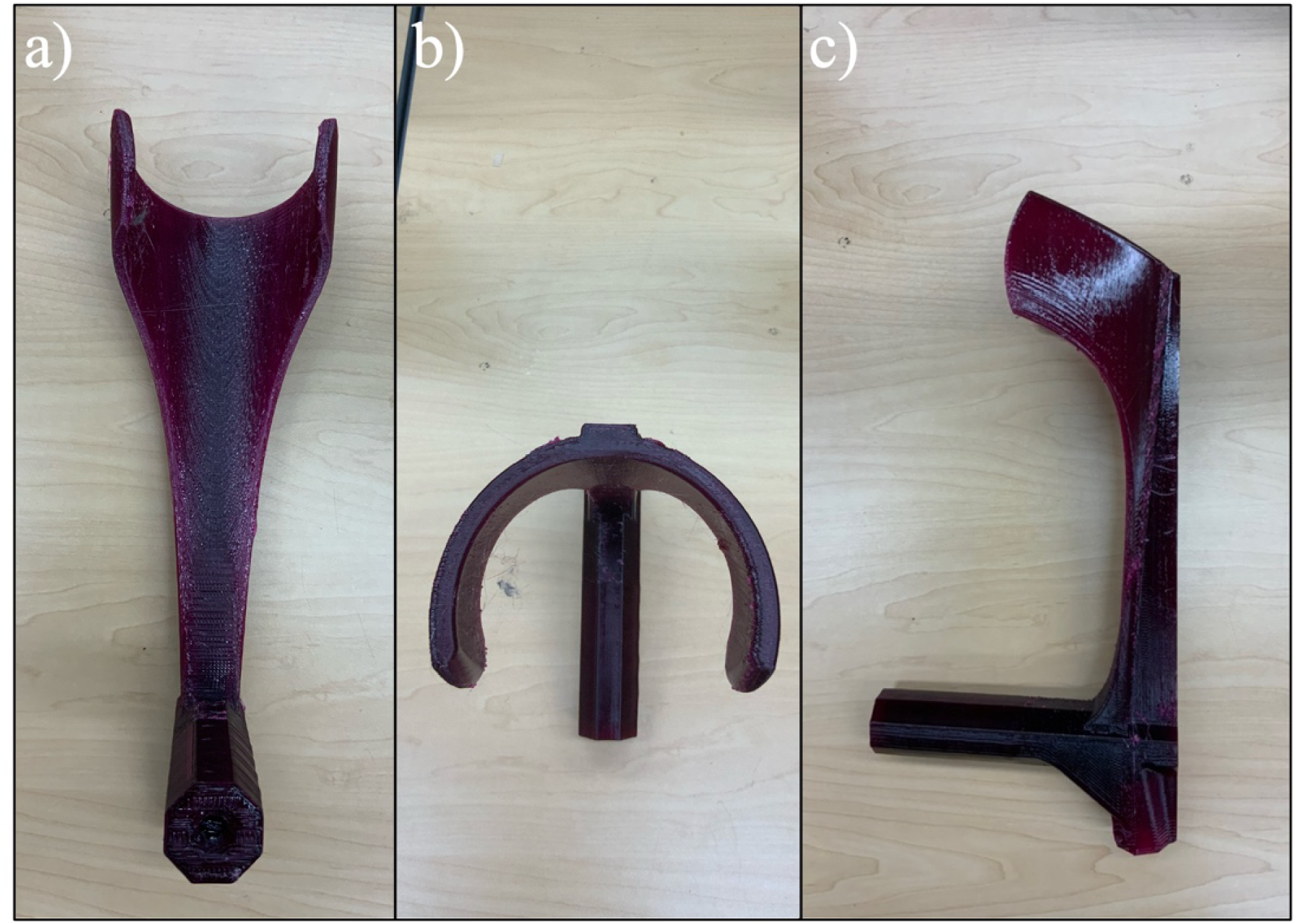
3D printed Forearm (PETG) a) front view, b) top view, c) side view.

**Figure 8.**
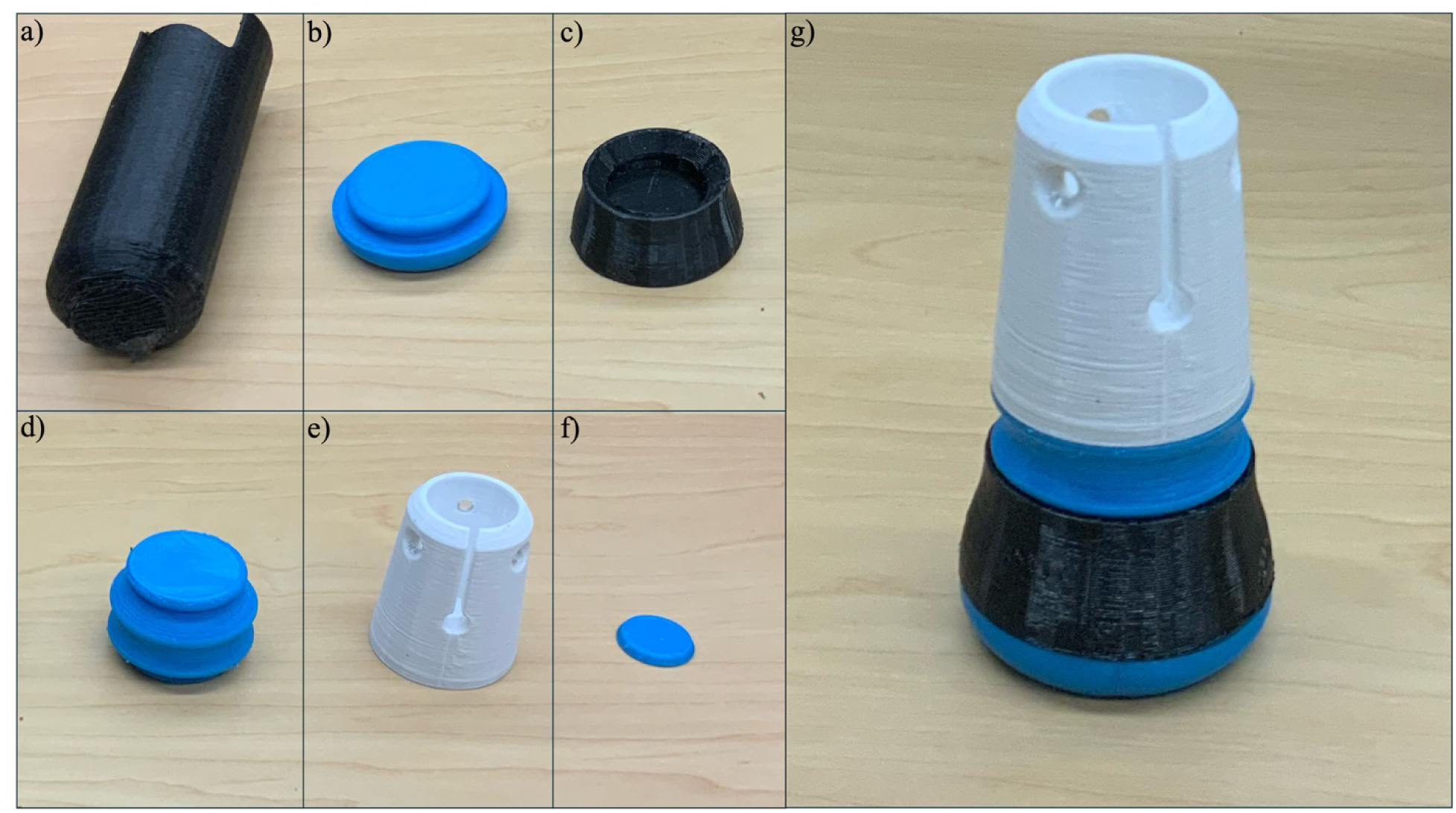
3D printed a) Handle Grip, b) Foot cushion, c) Foot, d) Foot living joint, e) Foot body, f) Washer, and g) Assembled Foot tip

**Figure 9.**
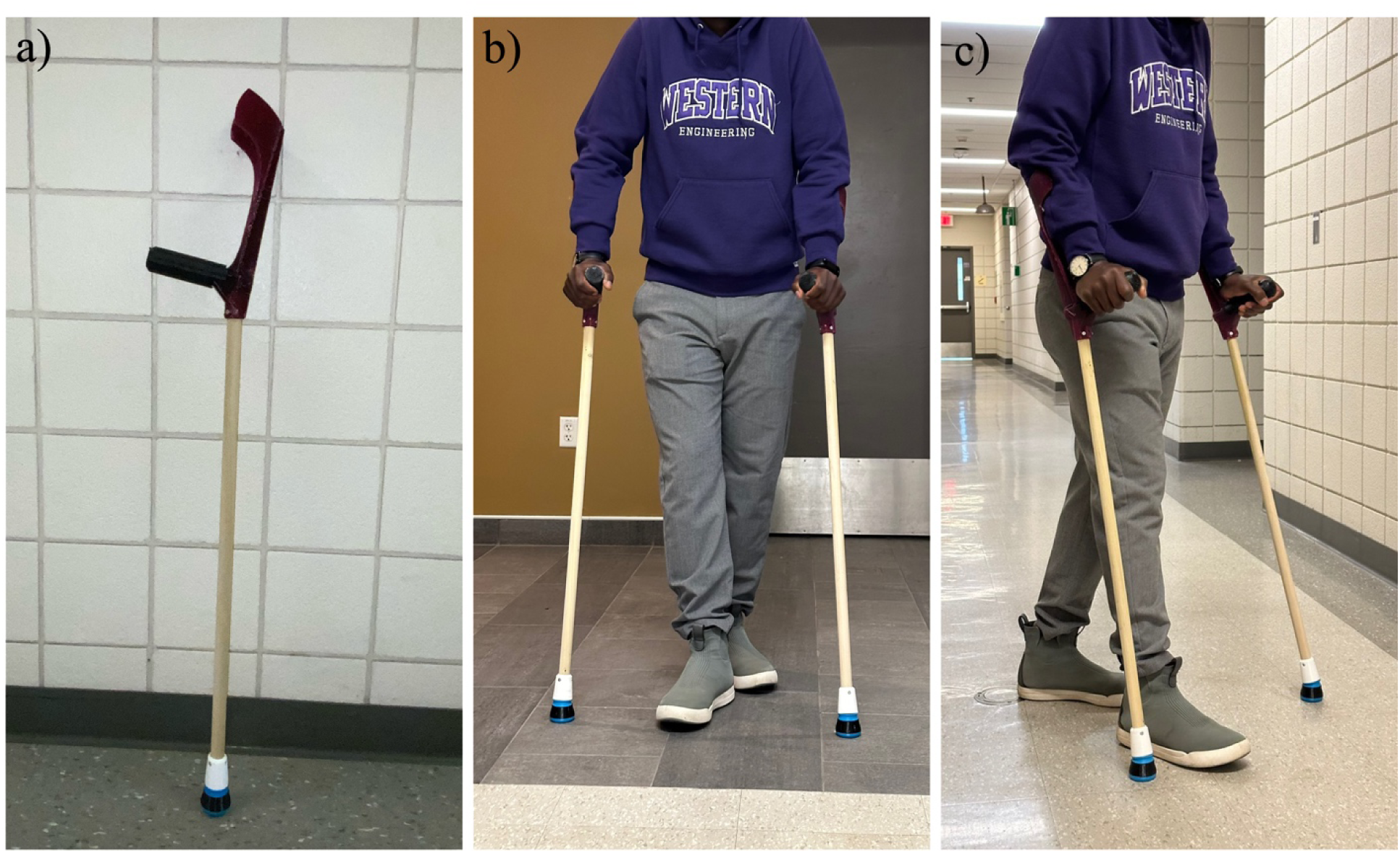
a) Assembled crutch, b and c) Two crutches in use

The load-displacement results of the five crutches are plotted in Figure 10. All crutches not only achieved the 1000N load prescribed by ISO 11334-1:2007 for the required duration of 10s, they proceeded to fail at loads more than 39% higher than the specified limit. The maximum compressive loads recorded for the five crutches tested were 1,852.8 N, 1,419.3 N, 1,459.6 N, 1,396.1 N, and 1,453.8 N, with an average of 1,516.32 ± 169.9 N. The failure loads were associated with vertical displacements that ranged from 29.5 mm to 37.9 mm, which result in an average displacement of 32.5 ± 3.2 mm. To determine the mass that the crutches can support, the compressive failure loads were converted to equivalent mass values using Newton’s second law:

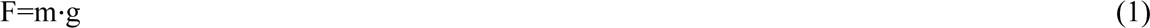

**Figure 10.**
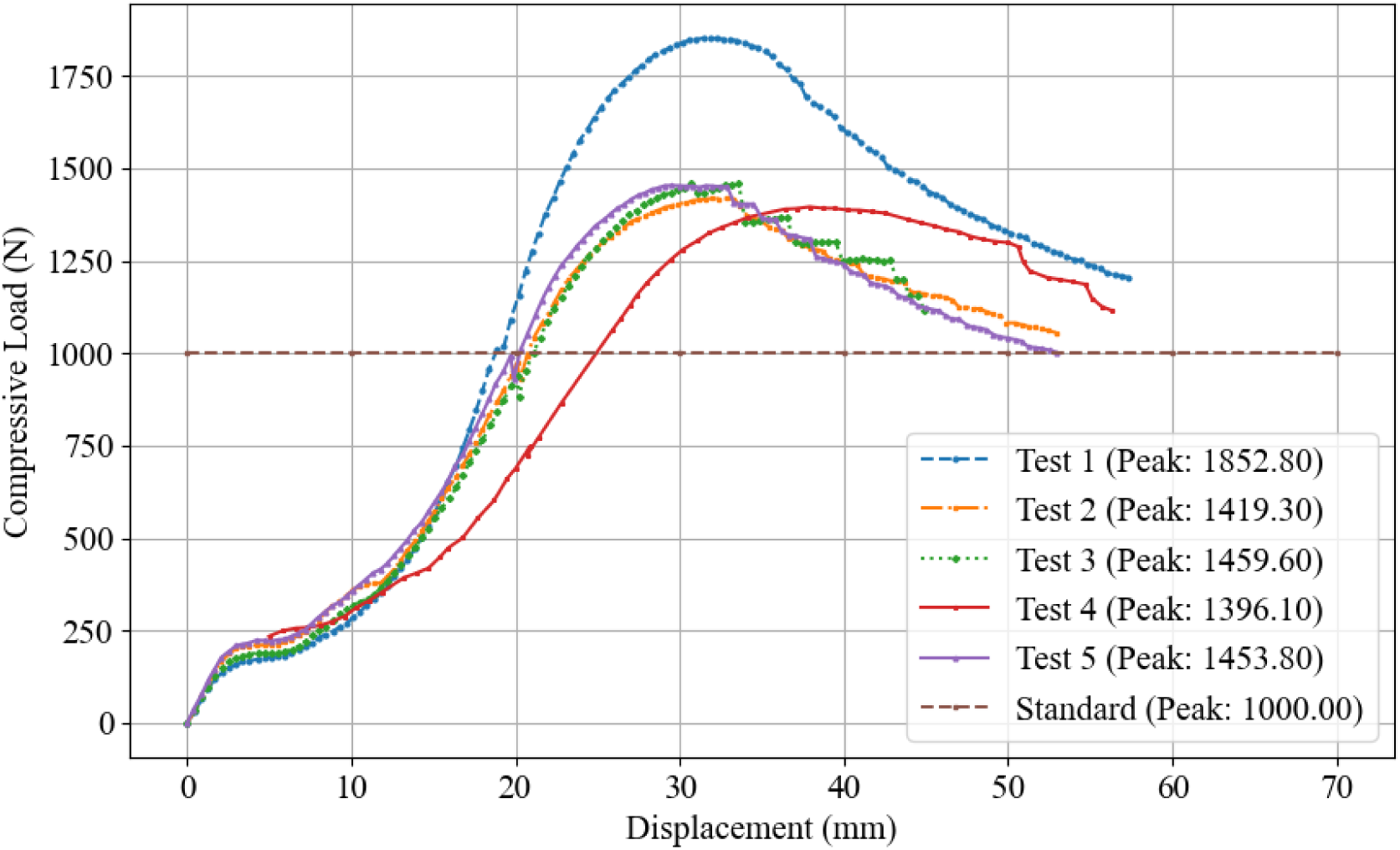
Compressive load vs displacement for the one-piece forearm crutch.

where F is the force (N), m is the mass (kg), and g is the acceleration due to gravity (9.81 m/s²). The corresponding mass values were calculated as 188.9 kg (416.4 lbs), 144.7 kg (319 lbs), 148.8 kg (320 lbs), 142.3 kg (313.7 lbs), and 148.2 kg (326.7 lbs), respectively, with an average of 154.6 ± 17.3 kg (340.8 ± 38.1 lbs). It is important to note that ISO 11334-1:2007 also suggests additional consideration for dynamic loading conditions, such as walking or climbing stairs. Forearm crutches are generally used two at a time with both in contact with the ground simultaneously, therefore the mass of a person using them can be divided by two. To ensure safety under these dynamic conditions, however, a safety factor of 2 is recommended. These two values cancel. Thus, using a safety factor of 2, the body weights the crutches can safely accommodate for dynamic use ranges from 142.3 kg (313.7 lbs) to 188.9 kg (416.4 lbs), with an average of 154.6 ± 17.3 kg (340.8 ± 38.1 lbs). It should be noted that this is conservative because it is unusual for a user’s full bodyweight to be placed on the crutches. Typically, users will support some of their bodyweight with their legs as well - though this will vary depending on the individual’s capabilities. The results also show that the maximum load capacity exceeded the test threshold of 1,000 N, which confirms that the crutches are designed to withstand significantly greater loads than typically expected during normal use or by the average user. This indicates that the crutches not only meet, but also surpass the required safety standards for effective performance according to these criteria. The videos of the tests are included in the Open Science Framework repository for the project [44].

When assessing the failure modes for each of the crutches, it was observed that four crutches failed at the dowel dure to bending (Test 1, Test 2, Test 4, and Test 5), while the 3D printed parts remained intact. The remaining study (Test 3, the orange filament) failed at the dowel- crutch attachment point. This failure point could indicate a point where wall thickness could be increased in future crutch designs to improve strength; particularly if stronger woods can be used for the dowel component. These failure modes are summarized below in Figure 11.

**Figure 11.**
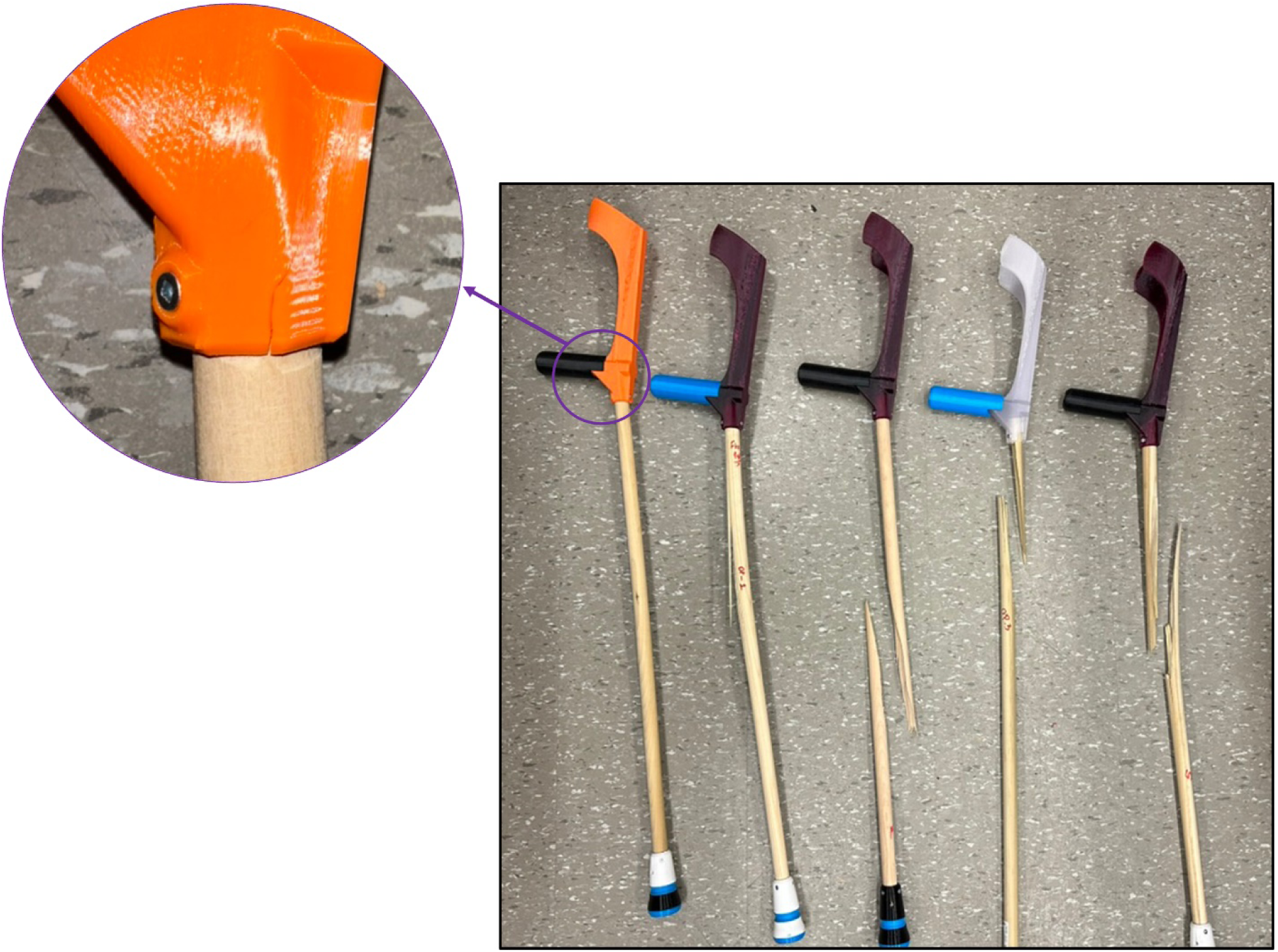
failure points for each of the 2 types of failure.

Table 4 summarizes the results of the 3D printed parts including mass, cost and manufacturing time. These results indicate that the most substantial time and material demand for this fabrication process is on the one-piece forearm component which takes approximately 25.4 hours to fabricate.

**Table 4.** 3D printed parts weight, price, and time breakdown.

| **PETG Parts \*** | **Quantity** | **Weight/Part (g)** | **Total Weight (g)** | **Cost (CAD)** | **Time** |
| --- | --- | --- | --- | --- | --- |
| One-piece forearm | 1 | 424 | 424 | 10.26 | 1 d 1 h 26 min |
| Ankle body | 1 | 33 | 33 | 0.79 | 2 h 25 min |
| Foot | 1 | 24 | 24 | 0.58 | 2 h 24 min |
| Total of PETG |  | 481 | 481 | 15.03 |  |
| **TPU Parts \*\*** |  |  |  |  |  |
| Handle Grip | 1 | 60 | 60 | 1.45 | 6 h 43 min |
| Washer | 1 | 1 | 1 | 0.02 | 00 h 06 min |
| Foot cushion | 1 | 32 | 32 | 0.77 | 2 h 49 min |
| Foot living joint | 1 | 33 | 33 | 0.79 | 2 h 57 min |
| Total of TPU |  | 126 | 126 | 8.36 |  |
| Total of PETG and TPU |  | 607 | 607 | 23.40 |  |
| *Cost is calculated based on a 1 kg Polymaker PETG Filament spool priced at CAD \$27.99 CAD + HST. ** Cost is calculated based on a 0.5 kg NinjaFlex TPU 85A Filament spool priced at CAD \$52.77+ HST. | | | | | |

The costs of the open-source crutch are summarized in Table 5. The total material cost of the open-source crutch is CAD $35.38. The 3D printed plastic represents approximately 66% of the total cost.

**Table 5.**
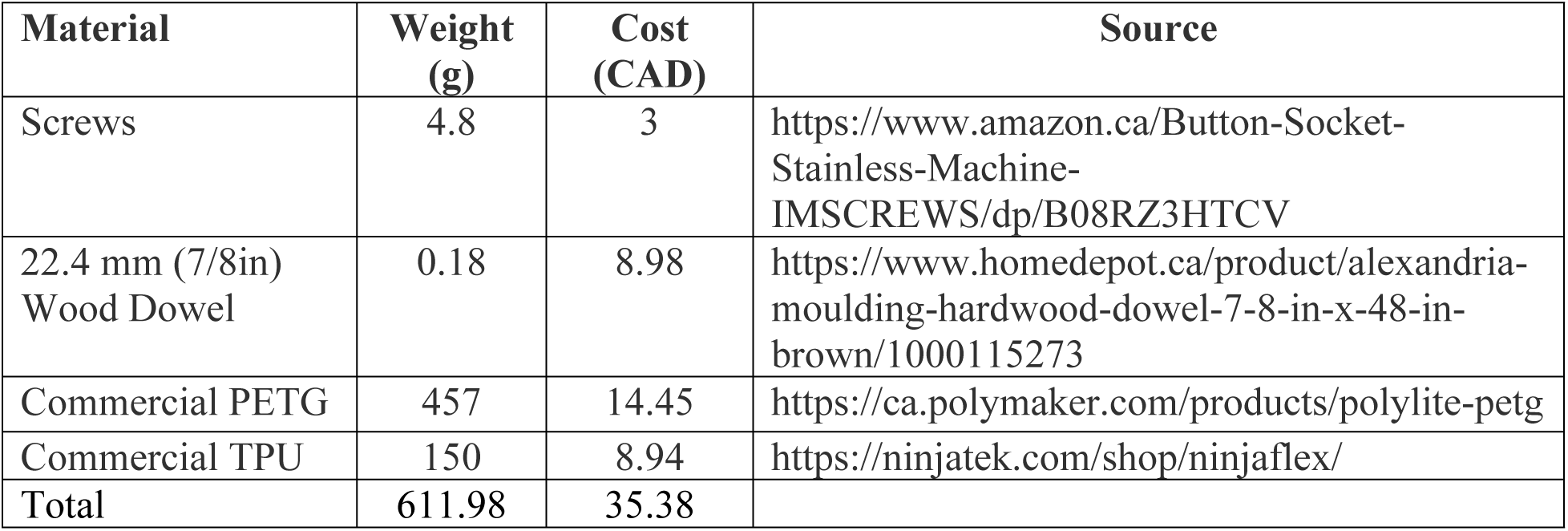
Cost calculation of the open-source crutch.

The open-source crutch offers significant advantages in terms of load capacity, materials, weight, cost, and customization. During the mechanical test, the open-source crutch demonstrated an average capacity per crutch of 1,516.3 ± 169.9 N or 154.6 ± 17.3 kg (340.8 ± 38.1 lbs), which exceeds the ISO 11334-1:2007 standard of 1,000 N or 101.9 kg (224.65 lbs), and commercial peers with the reported capacity of 113.4 kg (250 lbs) [45]. This means the open source crutches are capable of safely supporting users with body weights higher than the average weight of Canadian men (86.4 kg) and women (72.1 kg) with a safety factor of 1.78 and 2.17, respectively [46]. It should be noted that since each crutch is supporting half of the total weight, the safety factor effectively doubles when two crutches are utilized. Therefore, the effective safety factor becomes 3.56 for men and 4.34 for women. This strength makes the open-source crutch suitable for a broad range of users. A plot of the single crutch capacity relative to the weight distributions of Canadian men and women is provided for consideration in Figure 12.

**Figure 12.**
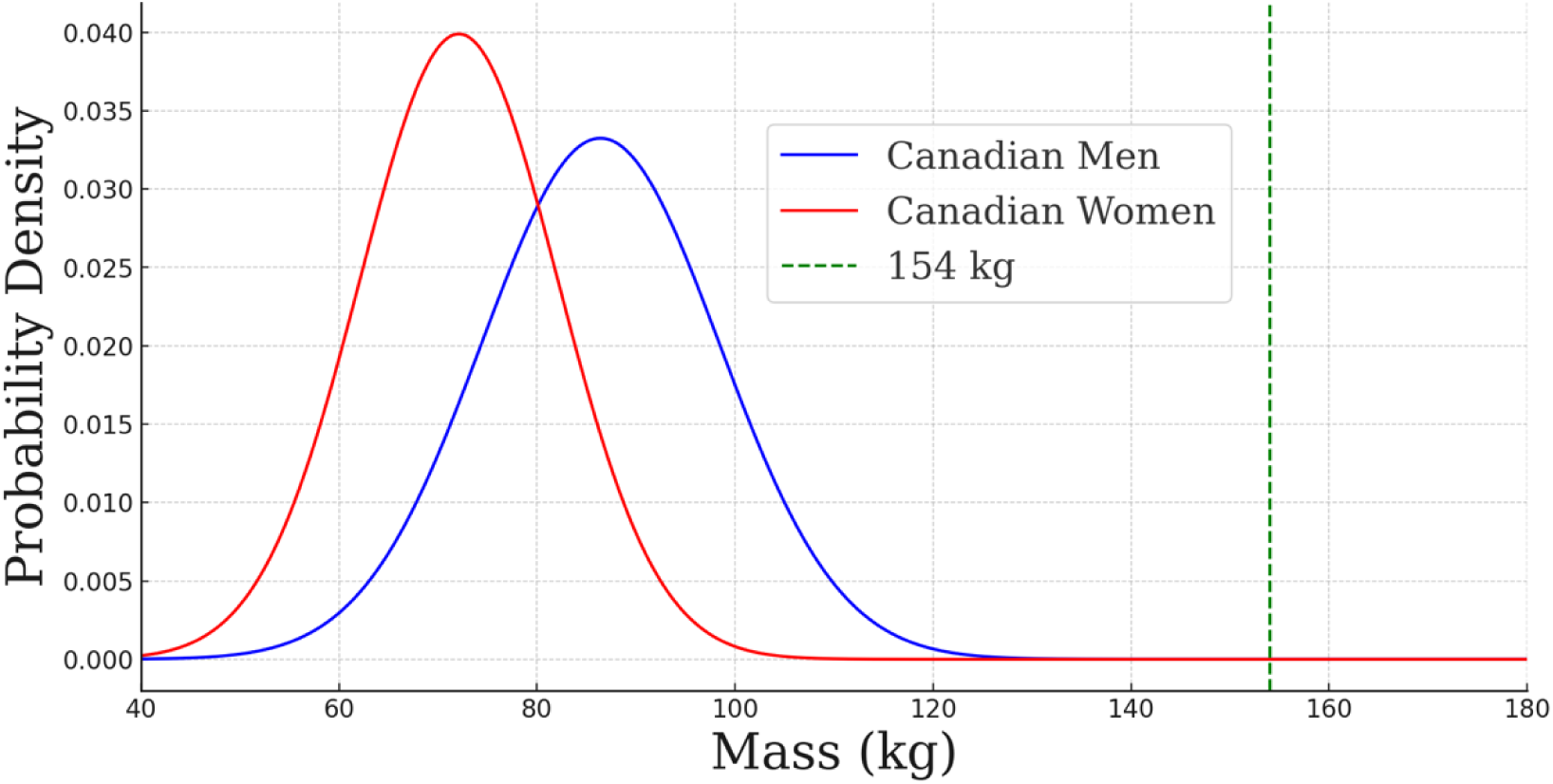
Gaussian distribution of masses of Canadian men and women in comparison to the average weight a pair of crutches can support under dynamic conditions (154 kg) using a safety factor of 2.

Most commercial crutches are typically made of aluminum and plastic. Although these materials provide durability, they also result in heavier products. For example, a similar commercial forearm crutch made of aluminum and plastic weighs approximately 2.27 kg [47]. In contrast, the open-source crutch weighs just 0.612 kg, which offers a weight reduction of approximately 73%, which promotes less labour-intensive use for long durations and promotes ease of crutch transport.

Using 3D printed parts in the open-source crutch provides additional benefits, such as flexibility, weight reduction, and the ability to customize the crutch to the user’s specific needs. This is in direct contrast with commercial crutches, which are generally mass-produced with limited options for personalization. Additionally, the use of TPU thermoplastic feet and handle covers improves shock absorption, which offers comfort during extended periods of use.

From an economic perspective, the open-source crutch is highly cost-effective, with a production cost of just CAD $35.38, which is lower than the commercial alternatives that offer similar performance (see Table 1; it is less expensive than forearm crutches, but not cheap underarm crutches). One way to decrease the cost of the crutch below that of all commercial alternatives is to use distributed recycling and additive manufacturing (DRAM) [48,49]. In this model, local waste plastic could be sourced individually [50] or at a community scale [51] and converted into 3D printing feedstocks [52]. DRAM manufacturing can reduce the cost of 3D printing materials to USD$0.025/kg [53]. As of now, the amount of 3D printing materials used is 607 g which results in a cost of approximately CAD $23.40 with commercial filament (USD $16.85, based on an exchange rate of 1 CAD$ = 0.72 USD$). With the implementation of DRAM manufacturing, the material cost would be USD$0.015 (CAD$0.021). This reduction would correspond to an overall decrease in the cost of the crutch equivalent to approximately CAD$12.98. To effectively use these materials, further testing would be needed to ensure the mechanical properties could be maintained with recycled plastic despite the mechanical recycling process reducing material strength.

Despite the benefits of the one-piece forearm crutch, it does have some limitations. One of the main drawbacks is the high amount of plastic material required for 3D printing. While 3D printing allows for customization and design flexibility, the large volume of this forearm crutch and high infill demands a lot of material, which increases production time. Although, 3D printing has an excellent ecological balance sheet, the greater the materials used, the greater the environmental impact [54,55]. Additionally, the forearm piece is quite large, which makes it challenging to print on smaller lower-cost desktop printers. As it is, a printer with a print bed of 300mm x 300mm was used to print the largest component. Another limitation is the lack of on-the-go height adjustability. While the dowel can be sized to fit the user, the design does not allow the users to easily modify the height while using it without replacing the dowel. It should also be noted that in general, as seen in the results, the wood was the weakest component. In one case, however, using a different color of filament caused the 3D printed component to fail first. It is well established in the literature that different colors of filaments have different strengths [56].

This variance, however, still demonstrated large variations in safety factors within the design.

There are several areas of future work. First, the amount of printing could be reduced by making the one-piece forearm out of two pieces. This would involve printing the handlebar and cuff as separate components, which could then be connected by a secondary wooden dowel. By doing so, the amount of plastic used would be reduced, the parts would be small enough to fit on a wider range of 3D printers (e.g. very small desktop printers), and the overall print cycle time would be reduced.

Another future direction could involve fabricating the entire crutch as a single 3D printed piece. Although this would require more filament and specialized 3D printers, it would allow for greater customization to meet the specific needs of users. Additionally, the development of a crutch with an adjustable height is suggested. This would enable height changes during use, making the crutch suitable for multiple users and would allow crutch manufacturers to stock pre-assembled crutches that allow for minor variations to suit users at the time of purchase.

## Conclusions

In conclusion, the open-source forearm crutch developed in this study not only surpasses commercially available alternatives in terms of load capacity, weight, and customization, but also offers a highly cost-effective solution. With an average load capacity of 1,516.3 ± 169.9 N, a lightweight design (0.612 kg), and a total production cost of only CAD$ 35.31 using virgin plastic, it is accessible to a broad range of individuals. This is particularly important for people with disabilities, who often face higher unemployment rates and financial challenges.

Additionally, 3D printing technology allows for distributed production, customization and enhanced comfort. While the crutch has proven effective, future improvements could address limitations such as the high plastic usage, print time and the fixed height of the design.

## Acknowledgements

This work was supported by the Thompson Innovation Fund and the Frugal Biomedical Program.

## 4. Data availability

All data and designs are available: https://osf.io/4b89v/ OSHWA Certification UID is: CA000058

